# Cardiovascular Risk Factors are Associated with Cerebrovascular Reactivity in Young Patients

**DOI:** 10.1101/2024.06.06.24308569

**Authors:** Jaskanwal DS Sara, Jay J. Pillai, Lilach O Lerman, Amir Lerman, Kirk Welker

## Abstract

**Introduction:** Endothelial dysfunction represents the earliest detectable stage of atherosclerosis, is associated with an increased risk of cardiovascular events, and predicts cardiovascular disease (CVD) more effectively than traditional cardiovascular risk factors. Cerebrovascular reactivity (CVR) provides an index of endothelial function in the brain. Poor CVR is associated with stroke, cerebral small vessel disease, dementia, and even coronary artery disease. Traditional CVD risk factors are associated with low CVR in patients with known CVD and in older cohorts. However, the relationship between cardiovascular risk profile and reduced CVR in young adults who do not yet have CVD is uncertain. We hypothesized that in young adults undergoing routine clinical fMRI examinations for non-vascular disease low CVR measures would be associated with increased cardiovascular risk factors.

**Methods:** This cross-sectional study included adults who underwent a clinically indicated 3-Tesla fMRI scan of the brain for mapping of eloquent cortex including a “breath-hold task” as an imaging quality control measure. Individuals with intracranial masses and those with baseline CVD were excluded. The task consisted of 5½, 20-second blocks of normal breathing interspersed with 20-second blocks of continuous breath holding. In breath hold fMRI scans, a voxel-wise comparison of brain T2 signal to an expected hemodynamic response curve is used to generate maps of voxel-wise t-statistics, indicating the probability that blood flow within a specific voxel had increased in response to changes in blood carbon dioxide levels. Using an axial slice 8 mm superior to the corpus callosum, a mean cerebral t-statistic was calculated for the slice as a comparative global measure of CVR in each patient. We retrospectively reviewed the charts of all individuals to characterize their clinical profile at the time of the fMRI. Based on the distribution of mean t-statistic values the sample was divided into two groups: high t-statistic (“normal reactivity”) and low t-statistic value (“abnormal reactivity”). The distribution of cardiovascular risk factors was then compared across groups.

**Results:** Between January 2014 and December 2023, 76 individuals underwent brain fMRI employing a “breath hold task” with suitable image quality for the current analysis (mean ± SD age, 35.46 ± 12.09 yrs.; 31.6% female). Mean ± SD global CVR T-statistic was 3.97 ± 1.62. Low CVR was defined as a mean T-statistic ≤4.2 (n=44, 57.9%). Individuals with abnormal CVR were older (age: 45.1 ± 10.3 vs. 27.0 ± 3.4 yrs., p<0.001), had a higher frequency of hypertension (31.8% vs. 14.3%, p=0.0069) and hyperlipidemia (18.2% vs. 3.1%, p=0.0449), and had higher systolic (123.5 ± 13.2 vs. 116.9 ± 12.2 mmHg, p=0.0282) and diastolic blood pressures (77.9 ± 11.8 vs. 72.2 ± 8.9, p=0.0141). Age, systolic blood pressure and hyperlipidemia were significantly associated with abnormal CVR in univariable and multivariable analyses (age, increase by 10 years OR: 2.00, 95% CI 1.40 – 2.78, p=0.0078; hyperlipidemia OR: 8.54, 95% CI 1.07 – 184.9, p=0.0049, and systolic blood pressure (OR for an increase in 10 mmHg: 1.57, 95% CI 1.10 – 2.10, p=0.0084).

**Conclusion:** Traditional cardiovascular risk factors are significantly associated with abnormal CVR in young adults without baseline CVD or cerebrovascular disease undergoing fMRI for reasons related to a diagnosis of epilepsy. Thus, CVR using fMRI could provide an integrated index of the collective burden of cardiovascular risk factors that could form a therapeutic target to prevent cardiovascular events.

## INTRODUCTION

Cerebral blood flow (CBF) autoregulation is a critical feature of the cerebral vasculature to ensure that the metabolic demands of the brain are met ^1^. Cerebrovascular reactivity (CVR) refers to dynamic changes in vascular tone in response to alterations in systemic blood pressure – mechanoregulation - and local carbon dioxide (CO2) concentrations - chemoregulation. ^1^. Unlike mechanoregulation, dynamic changes in CBF in response to CO2 represent an endothelium-dependent process affected by the production and availability of local nitric-oxide (NO) ^2–4^. Thus, endothelial-dependent cerebral vasomotion can be assessed by measuring CVR in response to CO2 by means of inhaling CO2-enriched gas ^5^ or through voluntary modulations of breathing pattern such as hyperventilation, or as is most commonly used, breath-holding ^5^. Such a protocol coupled with functional magnetic resonance imaging (fMRI) provides for a noninvasive imaging technique to quantify CVR in all brain tissues that does not involve ionizing radiation ^6^.

Low CVR is associated with cerebral steno-occlusive disease ^7,8^, small vessel disease ^9^, stroke ^10,11^, coronary artery disease (CAD) ^12^, Alzheimer’s disease, vascular dementia, and cognitive impairment ^13^. Lower CVR has also been detected in older age groups ^14,15^, in patients with obstructive sleep apnea ^16^, and in those with traditional cardiovascular risk factors such as hypertension, diabetes mellitus, and hypercholesterolemia ^17–19^. Vascular risk factors play a role in reducing the bioavailability and activity of NO ^20^ from the endothelium of vascular beds throughout the body, including within the brain ^19^. Indeed, endothelial dysfunction is the unique “response to injury” of the vasculature to a variety of risk factors ^21,22^, is the first stage of atherosclerosis, and is associated with an increased risk of ischemic cardiac events and stroke ^23–28^. Individuals with minimal traditional cardiovascular risk factors who have peripheral vascular endothelial dysfunction have a greater incidence of cardiovascular events and death compared to those with normal endothelial function ^29–31^ suggesting that endothelial dysfunction may be more effective at predicting cardiovascular disease (CVD) in seemingly low-risk populations. Thus, endothelial-dependent CVR measured in response to changes in CO2 concentration, may similarly be useful in estimating cardiovascular risk in groups traditionally thought to be at low risk.

Most studies that have evaluated the relationship between CVR and cardiovascular risk factors have included patients with known CVD ^7,8,10–12^ or older age groups ^17,18^ who would therefore have an elevated baseline risk irrespective of traditional risk factors or examined few CVD risk factors ^17^. Thus, to eliminate the effect of age, the aim of the current study is to compare the cardiovascular risk profile of a population of young adults (average age <40 years) without established CVD or cerebrovascular disease based on their CVR.

## METHODS

This cross-sectional study was approved by the Mayo Clinic Institutional Review Board, and all study participants provided their informed consent. The STROBE guidelines for cross-sectional studies were adhered to when reporting this study. We included all adults aged ≥ 18 years who underwent a clinically indicated 3-Tesla fMRI scan of the brain with a “breath hold task” to assess CVR (see below) since 2014 for reasons related to a diagnosis of epilepsy. Individuals with epilepsy typically undergo this type of imaging to facilitate pre-operative planning for epilepsy-related surgery. The following were excluded from the study: i) individuals with intracranial masses/space-occupying lesions, ii) individuals with existing baseline CVD or cerebrovascular disease, iii) individuals whose breath hold fMRI scans were deemed to be of poor quality due to excessive head motion (defined as greater than 1.5 mm of translational motion during the study) or inability or marginal ability to perform the breath hold task and iv) those who declined to have their medical records reviewed for research purposes. We then retrospectively reviewed the charts of all included individuals to characterize their clinical profile with respect to cardiovascular risk factors.

### Acquiring and Processing Data on Cerebrovascular Reactivity

The participants’ breath hold fMRI scans were previously acquired using a visually prompted block paradigm. This task consisted of 20 second blocks of normal breathing alternating with 20 second blocks of continuous breath holding. The task consisted of 5 ½ epochs, beginning and ending with a normal breathing block. If subjects were unable to hold their breath for a full 20 seconds, they were instructed to hold their breath as long as possible during the breath hold blocks.

The participants’ fMRI breath hold brain scans were performed on a Siemens Prisma 3T MRI scanner (Siemens Healthineers, Erlangen, Germany) using a 64-channel head coil. The following MRI scan parameters were employed for the fMRI acquisition: axial EPI/GRE pulse sequence; TR/TE = 2500/30 ms; flip angle = 90 degrees; FOV = 24 cm x 24 cm; slice thickness = 4 mm; matrix = 64 x 64; volumes 92 (with 4 discarded acquisitions for equilibration purposes). Additionally, all subjects previously underwent T1 weighted structural imaging of the brain (pre- or post-gadolinium) for anatomic localization purposes with the following approximate MRI scan parameters: axial T1 MPRAGE pulse sequence; TR/TE = 2100/2.26; flip angle = 10 degrees; FOV 25 cm x 25 cm; slice thickness = 1 mm; matrix 256 x 256.

The previously acquired breath hold fMRI scans were analyzed using Prism Process and Prism View software (Prism Clinic Imaging, Elm Grove, WI ). The data analysis method employed in this FDA-approved, commercially available software is based on the non-FDA-approved Analysis of Functional NeuroImages (AFNI) software created by the NIH and routinely employed in neurocognitive research. Image preprocessing steps included realignment of individual slices by means of a 6-degree affine transformation, spatial blurring to remove high-frequency noise, and slice timing correction. This was followed by voxel-wise comparison of signal to the expected breath hold task hemodynamic response function to generate voxel-wise statistical maps indicating the probability that an individual voxel’s blood oxygen level dependent (BOLD) signal change reflects a response to the blood carbon dioxide stimulus. The hemodynamic response function employed was based on prior research by Birn and colleagues (32). They demonstrated that, on average, cerebral BOLD signal peaks 30 seconds after the onset of a 20 second breath hold and that BOLD signal returns to baseline after another 20 seconds ^32^.

To measure relative CVR on the breath hold fMRI scans, for each subject, a brain mask was first obtained by skull stripping the T1 MPRAGE image with the Prism Process software. This mask was applied to the breath hold fMRI maps to eliminate extracranial image noise. To eliminate both artifacts from skull base susceptibility and the contribution of ventricular signal, an image slice 8 mm thick superior to the corpus callosum was selected. The cerebral cortex at this location is supplied medially by the anterior cerebral artery and laterally by the middle cerebral artery. The mean voxelwise t-value occurring in this slice in response to breath holding was calculated and used as a comparative measure of CVR. Negative BOLD signal occurring within the slice was excluded from the analysis as this generally represents vascular steal from white matter in response to the breath hold stimulus. For each patient we derived a mean cerebral t-statistic from the slice of interest as a ‘global index’ of CVR.

### Patient Information

Data was collected on conventional cardiovascular risk factors including hypertension, diabetes mellitus, hyperlipidemia, smoking status and body mass index (BMI); hematological and biochemical parameters including hemoglobin, white cell count, platelet count, creatinine, fasting blood glucose, total cholesterol, low density lipoprotein (LDL), high density lipoprotein (HDL), triglycerides, thyroid stimulating hormone (TSH), B12 and folate; and medication use including lipid-lowering therapy and selective serotonin receptor uptake inhibitors. Smoking was categorized as a history of current and former smoking or never smoking; hyperlipidemia was defined by a documented history of hyperlipidemia, treatment with lipid-lowering therapy, a LDL cholesterol level above the target (<130 mg/dL for low risk patients, < 100 mg/dL for moderate-high risk patients, <70 mg/dL for very high risk, and < 55 mg/dL for extreme high risk patients based on 10-year atherosclerotic cardiovascular disease risk), high-density lipoprotein cholesterol < 40 mg/dL in men or < 50 mg/dL in women, or triglycerides > 150 mg/dL. Type 2 diabetes mellitus was defined as a documented history of or treatment for type 2 diabetes, or an HbA1c of >6.5, if available. Hypertension was defined as a documented history of or treatment of the condition. All blood test results included in this study are based on blood samples obtained within 1 month of the index study. Information was also collected from ECGs obtained within 1 month of the index study including PR interval, QRS duration and corrected QT interval, as well as the systolic and diastolic blood pressure and heart rate obtained on the day of the study ^33–37^. We also collected information on the presence of a diagnosis of obstructive sleep apnea, major depressive disorder or anxiety disorder as documented in the patient’s medical history.

### Statistical analyses

There are currently no available normative data with respect to normal and abnormal CVR T-statistic values. Thus for the current study, we evaluated the distribution of slice-wise mean T-statistic values across our study population and used the average value as measure to divide our sample into two groups: those with high T-statistic values (“normal CVR”) and those with low T-statistic values (“abnormal CVR”). We then compared baseline clinical characteristics across the groups and presented these in Table 1. Next the CVR T-statistic as a continuous variable was compared across various cardiovascular risk factors that were grouped in a binary fashion (i.e., sex: men vs. women; hypertension: yes vs. no; diabetes mellitus: yes vs. no etc.). The Pearson’s correlation was then computed for the CVR T-statistic with continuous cardiovascular risk factors such as age, BMI, systolic blood pressure etc. Next, we estimated the odds ratio of having ‘abnormal CVR (low T-statistic values) as a categorical independent variable for various cardiovascular risk factors in univariable and multivariable analyses. Continuous variables are presented as a mean (standard deviation), and categorical variables are presented as frequencies (percentages). Differences between groups were analyzed using Student’s T test for continuous variables and Pearson’s chi-squared test for proportions. A p-value less than 0.05 was considered significant and all statistical analyses will be performed using BlueSky Statistics software 2022.

**TABLE 1.** Summary of Baseline Clinical Characteristics of Patients with Normal and Abnormal Cerebral Vasoreactivity.

|  | **Normal Cerebral**<br>**Vasoreactivity**<br>**N=32 (42.1%)** | **Abnormal Cerebral**<br>**Vasoreactivity**<br>**N=44 (57.9%)** | *P Value* |
| --- | --- | --- | --- |
| Age, years (SD) | 27.0 (3.4) | 45.1 (10.3) | <0.001* |
| Female, n (%) | 14 (43.8%) | 14 (31.8) | 0.2870 |
| BMI, kg/m<sup>2</sup> (SD) | 26.6 (5.5) | 27.7 (6.5) | 0.4243 |
| Hypertension, n (%) | 2 (14.3) | 14 (31.8) | 0.0069* |
| Diabetes Mellitus, n (%) | 0 (0.0) | 7 (6.8) | 0.1318 |
| Hyperlipidemia, n (%) | 1 (3.1) | 8 (18.2) | 0.0449* |
| Smoker (current or former), n (%) | 8 (25.0) | 18 (40.9) | 0.1489 |
| Obstructive Sleep Apnea, n (%) | 4 (12.5) | 14 (31.8) | 0.0505* |
| Major Depressive Disorder, n (%) | 18 (56.3) | 23 (52.3) | 0.7313 |
| Anxiety Disorder, n (%) | 15 (46.9) | 22 (50.0) | 0.7878 |
| Hemoglobin, mg/dL (SD) | 14.3 (1.6) | 14.2 (1.3) | 0.7558 |
| Leukocytes, x 10<sup>9</sup>/L (SD) | 6.5 (2.5) | 11.0 (2.1) | 0.4385 |
| Platelets, x 10<sup>9</sup>/L (SD) | 240.2 (61.5) | 230.9 (66.3) | 0.5463 |
| Creatinine, mg/dL (SD) | 0.9 (0.2) | 0.9 (0.2) | 0.8864 |
| eGFR, mL/min/1.73m<sup>2</sup> (SD) | 60.3 (1.6) | 60.0 (0.0) | 0.2360 |
| Glucose, mg/dL (SD) | 103.1 (19.0) | 104.1 (19.0) | 0.8359 |
| Total Cholesterol, mg/dL (SD) | 178.3 (41.8) | 192.6 (37.2) | 0.3546 |
| LDL Cholesterol, mg/dL (SD) | 102.7 (35.9) | 112.6 (30.5) | 0.4378 |
| HDL Cholesterol, mg/dL (SD) | 56.8 (15.0) | 57.8 (18.7) | 0.8817 |
| Triglycerides, mg (dL) (SD) | 99.2 (21.1) | 113.5 (75.6) | 0.6208 |
| Folate, ng/mL (SD) | 15.6 (6.3) | 14.3 (4.8) | 0.7655 |
| Vitamin B12, pg/mL (SD) | 608.5 (712.1) | 406.4 (195.0) | 0.3744 |
| TSH, mU/ml (SD) | 4.5 (3.7) | 2.7 (2.0) | 0.0425* |
| PR Interval, ms (SD) | 147.1 (32.9) | 155.2 (21.3) | 0.2683 |
| QRS Interval, ms (SD) | 92.6 (10.4) | 95.6 (10.8) | 0.3043 |
| Corrected QT Interval, ms (SD) | 420.2 (29.1) | 422.9 (21.0) | 0.6913 |
| Heart Rate, bpm (SD) | 73.1 (11.8) | 75.9 (12.2) | 0.3259 |
| Systolic Blood Pressure, mmHg (SD) | 116.9 (12.2) | 123.5 (13.2) | 0.0282* |
| Diastolic Blood Pressure, mmHg (SD) | 72.2 (8.9) | 77.9 (11.8) | 0.0141* |
| Aspirin Use, n (%) | 0 (0.0) | 2 (4.5) | 0.2216 |
| ACE-inhibitor/ARB Use, n (%) | 0 (0.0) | 3 (6.8) | 0.1318 |
| Bet Blocker Use, n (%) | 1 (3.1) | 6 (13.6) | 0.1177 |
| Calcium Channel Blocker Use, n (%) | 0 (0.0) | 3 (6.8) | 0.1318 |
| Thiazide or Loop Diuretic or Spironolactone Use, n (%) | 0 (0.0) | 5 (11.4%) | 0.0485* |
| Statin Use, n (%) | 0 (0.0) | 6 (13.6) | 0.0295* |
| Selective Serotonin Receptor Use, n (%) | 7 (21.9) | 12 (27.3) | 0.5916 |

Abbreviations – ACE-inhibitor: Angiotensin Converting Enzyme-Inhibitor; ARB: Angiotensin Receptor Blocker; BMI: Body Mass Index; eGFR: estimated Glomerular Filtration Rate; HDL: High Density Lipoprotein; LDL: Low Density Lipoprotein; TSH: Thyroid Stimulating Hormone

## RESULTS

### Sample Overview

Between January 2014 and December 2023, 87 patients (mean ± SD, age 35.21 ± 12.83 years; 35.6% female) underwent fMRI of the brain with a “breath hold task” for assessment of CVR. Seventy-six (87.4%) patients had fMRI images that were of sufficient quality to be included in this study (mean ± SD, age 35.46 ± 12.09 years; 31.6% female). Reasons for excluding patients from this analysis included excessive translational motion during the study and/or inability or only marginal ability to perform the breath-hold task appropriately as reported by the performing technicians. None of these patients had any baseline CVD including a history of CAD, myocardial infarction, peripheral arterial disease, stroke or transient ischemic attack. The mean ± SD global CVR T-statistic across the sample was 3.97 ± 1.62. The median value (Q1, Q3) was 3.90 (2.89, 5.04) and the mode was 4.62. Thus, we elected to divide our sample into two groups with a high T-statistic (‘normal CVR) and low T-statistic (‘abnormal CVR) using 4.2 as the cut-off given this value was approximately half-way between the median and the mode. Thirty two (42.1%) of individuals had a T-statistic > 4.2 and 44 (57.9%) had a T-statistic ≤ 4.2. **Table 1** summarizes the baseline characteristics of patients across both groups.

Individuals with abnormal CVR were on average older (age: 45.1 ± 10.3 vs. 27.0 ± 3.4 years, p<0.001), had a higher frequency of hypertension (31.8% vs. 14.3%, p=0.0069), hyperlipidemia (18.2% vs. 3.1%, p=0.0449) and obstructive sleep apnea (31.8% vs. 12.5%, p=0.0505) compared to those with normal CVR, albeit the latter difference was with borderline significance. Further, individuals with abnormal CVR had lower TSH values (2.7 ± 2.0 vs. 4.5 ± 3.7, p=0.0425) and higher systolic (123.5 ± 13.2 vs. 116.9 ± 12.2 mmHg, p=0.0282) and diastolic blood pressures (77.9 ± 11.8 vs. 72.2 ± 8.9, p=0.0141) compared to those with normal cerebrovascular reactivity. Last, individuals with abnormal CVR used diuretic medication (thiazides, loop diuretics or spironolactone: 11.4% vs. 0.0%, p=0.0485) and statins (13.6% vs. 0.0%, p=0.0295) more frequently than those with normal CVR. Otherwise, there were no other significant differences between groups concerning demographic, clinical, or laboratory variables or with respect to medication use **(Table 1).**

### Cardiovascular Risk Factors and Cerebrovascular Reactivity

**Table 2** demonstrates the relationship between cardiovascular risk factors dichotomized into groups (i.e., age > 33 or ≤ 33 years, male or female sex, etc.) and CVR T-statistic values as continuous variables. Individuals with an age > 33 years had on average lower T-statistic values compared to those aged ≤ 33 years (2.82 ± 0.9 vs. 5.39 ± 1.1, p<0.001) suggesting that older individuals have worse CVR. Individuals with hyperlipidemia showed a trend towards having on average lower mean t-statistic values compared to those without hyperlipidemia although this was not statistically significant (3.06 ± 1.3 vs. 4.09 ± 1.6, p=0.0720). There were no significant differences in t-statistic values across sexes, between those who were overweight (BMI > 25 kg/m^2^) compared to those with a normal BMI, or across those with versus those without hypertension, diabetes mellitus, obstructive sleep apnea, smoking history, depressive disorder or anxiety disorder.

**TABLE 2.** Cerebral Vasoreactivity Across Cardiovascular Risk Factors.

|  | Cerebral Vasoreactivity<br>T-statistic | *P* value |
| --- | --- | --- |
| ***Cardiovascular Variable*** |  |  |
| **Age** |  | <0.001* |
| > 33 yrs | 2.82 (0.9) |  |
| ≤ 33 yrs | 5.39 (1.1) |  |
| **Sex** |  | 0.2004 |
| Males | 3.79 (1.6) |  |
| Females | 4.28 (1.7) |  |
| **Hypertension** |  | 0.2146 |
| Yes | 3.52 (1.4) |  |
| No | 4.09 (1.7) |  |
| **Hyperlipidemia** |  | 0.0720 |
| Yes | 3.06 (1.3) |  |
| No | 4.09 (1.6) |  |
| **Diabetes Mellitus** |  | 0.4294 |
| Yes | 3.24 (1.1) |  |
| No | 4.00 (1.6) |  |
| **Obstructive Sleep Apnea** |  | 0.1081 |
| Yes | 3.43 (1.5) |  |
| No | 4.14 (1.6) |  |
| **Smoker (current or former)** |  | 0.8264 |
| Yes | 3.91 (1.5) |  |
| No | 4.00 (1.7) |  |
| **Body Mass Index** |  | 0.2856 |
| BMI > 25 kg/m<sup>2</sup> | 3.80 (1.6) |  |
| BMI ≤ 25 kg/m<sup>2</sup> | 4.20 (1.6) |  |
| **Major Depressive Disorder** |  | 0.7664 |
| Yes | 4.02 (1.5) |  |
| No | 3.91 (1.7) |  |
| **Anxiety Disorder** |  | 0.9552 |
| Yes | 3.96 (1.7) |  |
| No | 3.98 (1.5) |  |

The linear correlation between t-statistic values as a continuous variable and continuous cardiovascular risk factors was also assessed. The cerebrovascular reactivity t-statistic was inversely and significantly correlated with age (r: -0.4969, p<0.001). There was a trend to a significant correlation between the t-statistic and heart rate (r: -0.2163, p=0.0605). There were no significant correlations between the t-statistic and hemoglobin, white cell count, platelet count, blood glucose, TSH, total cholesterol, LDL cholesterol, HDL cholesterol, triglycerides, BMI, and systolic and diastolic blood pressure.

### Cardiovascular Risk Factors and Abnormal Cerebrovascular Reactivity – Univariable and Multivariable Analyses

**Table 3** demonstrates the association between cardiovascular risk factors and an abnormal CVR (t-statistic ≤ 4.2) in univariable analyses. Age, increase by 10 years (OR: 1.59, 95% CI 1.14 – 2.21, p=0.0149*), hypertension (OR: 7.00, 95% CI 1.76 – 47.08, p=0.0041), hyperlipidemia (OR: 6.89, 95% CI 1.17 – 131.46, p=0.0307) and obstructive sleep apnea (OR: 3.27, 95% CI 1.03 – 12.61, p=0.0442) were significantly associated with abnormal CVR. Systolic blood pressure was also associated with CVR (OR for an increase in systolic blood pressure of 10 mmHg: 1.40, 95% CI 1.10 – 1.90, p=0.0337). Other cardiovascular risk factors were not significantly associated with abnormal CVR.

**TABLE 3.** Univariable Analyses of the Relationship Between Cardiovascular Risk Factors and Abnormal Cerebral Vasoreactivity.

|  | **Odds**<br>**Ratio** | **95% Confidence**<br>**Interval** | *P value* |
| --- | --- | --- | --- |
| Age, increase by 10 years | 1.59 | 1.14 – 2.21 | 0.0149* |
| Male Sex | 1.09 | 0.43 – 2.76 | 0.8610 |
| Hypertension | 7.00 | 1.76 – 47.08 | 0.0041* |
| Hyperlipidemia | 6.89 | 1.17 – 131.46 | 0.0307* |
| Obstructive Sleep Apnea | 3.27 | 1.03 – 12.61 | 0.0442* |
| Smoker<br>(current or former) | 2.08 | 0.78 – 5.88 | 0.1450 |
| Overweight<br>(BMI > 25 kg/m<sup>2</sup>) | 1.75 | 0.70 – 4.47 | 0.2347 |
| Major Depressive Disorder | 0.85 | 0.34 – 2.13 | 0.7311 |
| Anxiety Disorder | 1.13 | 0.45 – 2.84 | 0.7878 |
| BMI, increase by 1 kg/m<sup>2</sup> | 1.03 | 0.96 – 1.12 | 0.4138 |
| Systolic Blood Pressure (increase by 10 mmHg) | 1.40 | 1.10 – 1.90 | 0.0337* |
| LDL Cholesterol (increase by 1 mg/dL) | 1.10 | 0.90 – 1.40 | 0.4124 |
| Triglycerides (increase by 1 mg/dL) | 1.00 | 0.90 – 1.20 | 0.6095 |

Abbreviations – BMI: Body Mass Index; LDL: Low Density Lipoprotein

**Table 4** demonstrates the association between cardiovascular risk factors and abnormal CVR in multivariable analyses adjusting for age, sex, systolic blood pressure, hyperlipidemia, BMI > 25 kg/m^2^ and obstructive sleep apnea. Age (for an increase in 10 years, OR: 2.00, 95% CI 1.40 – 2.78, p=0.0078), hyperlipidemia (OR: 8.54, 95% CI 1.07 – 184.9, p=0.0049), and systolic blood pressure (OR for an increase in systolic blood pressure of 10 mmHg: 1.57, 95% CI 1.10 – 2.10, p=0.0084) were significantly associated with abnormal CVR. Smoking status, BMI, and obstructive sleep apnea were not associated with abnormal CVR.

**TABLE 4.** Multivariable Analyses of the Relationship Between Cardiovascular Risk Factors and Abnormal Cerebral Vasoreactivity.

|  | **Odds Ratio** | **95% Confidence Interval** | *P value* |
| --- | --- | --- | --- |
| Age, increase by 10 years | 2.00 | 1.40 – 2.78 | 0.0078* |
| Systolic Blood Pressure (increase by 10 mmHg) | 1.57 | 1.10 – 2.10 | 0.0084* |
| Hyperlipidemia | 8.54 | 1.07 – 184.9 | 0.0049* |
| BMI, increase by 1 kg/m<sup>2</sup> | 0.95 | 0.85 – 1.05 | 0.8585 |
| Obstructive Sleep Apnea | 3.17 | 0.80 – 14.99 | 0.1162 |
| Smoker | 2.99 | 0.93 – 10.59 | 0.0754 |

Abbreviations – BMI: Body Mass Index

## DISCUSSION

In the current study we report the comprehensive cardiovascular profile of a cohort of young adults, with an average age < 40 years, without established CVD or cerebrovascular disease who undergo testing with a fMRI BOLD ‘breath-hold’ protocol to assess CVR. We demonstrate that traditional cardiovascular factors varied significantly between individuals with a higher “normal CVR” compared to a lower “abnormal CVR.” Specifically, older age, hypertension, hyperlipidemia, obstructive sleep apnea, and elevated blood pressure values were all more prevalent in those with a lower CVR. In univariable and multivariable analyses age, hyperlipidemia, and systolic blood pressure remained significantly associated with a lower CVR. Obstructive sleep apnea was associated with abnormal CVR in univariable analyses only, implying its relationship with abnormal CVR is confounded by its association with other cardiovascular risk factors such as blood pressure. Thus, the current study supports the concept that an abnormal or low CVR is independently associated with traditional cardiovascular risk factors even in young and apparently low-risk populations.

### Cerebrovascular Reactivity and Endothelial Dysfunction

Maintaining adequate CBF is vital to meeting the metabolic demands of the brain and is achieved by modifying cerebrovascular tone in response to a variety of clinical conditions. Dynamic changes in CVR may be modulated by local CO2 concentrations (chemoregulation) ^1^ in an endothelial-dependent manner affected by the availability of local NO ^3^. For example, in a primate model hypercapnia induced increases in CBF that were completely blocked when a NO synthase inhibitor was infused into the internal carotid artery ^2^. Further, tonic production of NO can be inhibited using N^G^-monomethyl-L-arginine (L-NMMA) whose effects on regional CBF are linearly correlated with arterial PCO2, and are reversed by L-arginine, a substrate for endothelial NO synthase ^2^. These findings imply a graded relationship between NO production and arterial PCO2. Moreover, systemic administration of L-arginine improved CVR in response to CO2 in individuals with impaired baseline CVR ^38^. Taken together, these findings suggest that the protocol used in the current study, namely CVR in response to breath-holding, evaluated cerebrovascular endothelial function.

Endothelial dysfunction is the earliest detectable stage of atherosclerosis and occurs as a “response to injury” of the vasculature to various clinical risk factors ^21,22^. Our findings suggest that traditional cardiovascular factors such as age, elevated blood pressure and hyperlipidemia may adversely affect endothelial health in the cerebral vasculature in a similar manner to the peripheral and coronary vasculature ^39^. This concept supports the notion that endothelial dysfunction is a systemic process and is inducible by similar risk factors across different vascular beds. Indeed, traditional cardiovascular risk factors reduce the bioavailability and activity of NO ^20^ from the vascular endothelium throughout the body including within the brain ^19^. In one study, diabetic and/or hypertensive patients with impaired peripheral endothelial function, measured using forearm blood flow in response to reactive hyperemia, exhibited reduced CO2-dependent CVR compared to controls who had normal peripheral endothelial function ^4^. These group differences were ameliorated following the administration of the NO donor nitroprusside ^4^. Moreover, endothelial dysfunction detected peripherally is associated with an increased risk of cardiovascular events and stroke ^23–28^. Population studies have shown that individuals with minimal traditional risk factors but who have peripheral vascular endothelial dysfunction have a higher incidence of cardiovascular events and death compared to those with normal endothelial function ^29–31^ suggesting that endothelial dysfunction may predict CVD more effectively in apparently low-risk populations. Along the same lines, we demonstrated that traditional cardiovascular risk factors were strongly associated with low CVR in adults who would traditionally be considered low-risk with an average age < 40 years and no baseline CVD. Thus, endothelial-dependent CVR measured using BOLD fMRI could provide for an integrated index of cardiovascular risk in young adults. At present prospective studies evaluating the implications of a low baseline CVR in patients without CVD are lacking but would be of great value in determining if CVR offers promise in predicting risk in such populations.

### Therapies Targeting Cerebrovascular Reactivity

Evaluating the clinical utility of measuring CVR as a marker of cardiovascular risk may be beneficial as pharmacologic and nonpharmacologic therapies improving CVR have been described in humans. L-arginine has been shown to improve CVR in response to CO2 in individuals with an abnormal baseline CVR in the context of cardiovascular risk factors such as hypertension but not in those with normal baseline CVR ^38^. L-arginine provides these effects as a necessary substrate to synthesize NO, but also through scavenging free radicals ^40^. L-arginine has also been shown to improve coronary endothelial function in hypercholesterolemic patients ^41^. Another study demonstrated that statin therapy resulted in improvements in CVR in individuals with cerebral small vessel disease, with more pronounced effects in those with the most severe impairments in CVR ^42^. Statin therapy has been shown to improve endothelial-dependent vasomotion in the coronary ^43,44^ and peripheral arteries ^45,46^. Statin-induced improvements in endothelial-dependent vasomotion are thought to be mediated by their lipid-lowering properties ^47^, and by enhancing NO synthase activity ^48^. Mild-moderate aerobic fitness training, as part of cardiac rehabilitation, has also been shown to be associated with improvements in regional CBF in individuals with CAD ^12^. Indeed, higher regional CBF has been demonstrated in aerobically trained adults compared to sedentary controls ^49^, and a positive correlation was shown between VO2 maximum and regional CBF in patients with CAD ^50^. Studies have also shown increased brain volume in specific regions, such as the medial prefrontal region, in response to exercise training ^51^, which may represent the cause or effect of changes in blood flow. Aerobic training also improves peripheral endothelial function in patients with CAD ^52,53^ and healthy subjects ^54^. Thus, the abovementioned treatment strategies appear to improve endothelial dysfunction across different vascular territories, and prospective studies evaluating their impact on CVR as a marker of cardiovascular risk will be of great value.

### Studies Evaluating Cerebrovascular Reactivity in Patients With or at Risk of Cardiovascular Disease

Studies have shown that a low CVR is associated with a variety of established CVDs including cerebral steno-occlusive disease ^7,8^, cerebral small vessel disease ^9^, CAD ^12^, and stroke ^10,55^. In fact, reduced CVR predicted peri-infarct T2 hyperintensities, greater infarct volume, and worse clinical outcomes in stroke patients ^11^. Studies have also demonstrated low CVR in Alzheimer’s disease, vascular dementia and cognitive impairment ^13^. In individuals without established CVD, a low CVR has also been detected in older age groups ^14,15^, in patients with obstructive sleep apnea ^16^, and in those with hypertension, diabetes mellitus and hypercholesterolemia ^9,17–19^. The largest of these included 541 participants using a similar breath-hold fMRI protocol as in our study and demonstrated reduced CVR in specific brain regions that underly the default-mode network in patients with prehypertension/hypertension ^17^. Differences were also observed in CVR between those with diabetes mellitus and hyperlipidemia compared to healthy controls although these were not significant ^17^. The average age of the population included in this study was 50 years, whereas the current study evaluated individuals with an average age of 35 years. Other studies included participants with average ages greater than 65 ^9,19^ or 70 ^18^ years. Thus, our study extends the findings of these previous studies by demonstrating that traditional cardiovascular risk factors such as hypertension and hyperlipidemia are independently associated with lower CVR even in young adults. Our study further extends previous studies in that we did not limit our analyses to outlining differences in categorical cardiovascular risk factors such as hypertension and diabetes mellitus etc. but instead comprehensively described the cardiovascular risk profile of our study participants by including the results of laboratory and ECG testing as well as cardiovascular medications.

Our study demonstrated that age was strongly associated with a low CVR in a sample with a mean age of 35 years. This highlights the fact that aging is a continuous process and that even young adults are at risk of a low CVR independent of other traditional risk factors and may thus benefit from early screening. Further, our findings underscore the importance of hypertension and blood pressure as risk factors for low CVR, which were also consistently demonstrated in previous studies in older populations ^9,17–19^. Obstructive sleep apnea was associated with low CVR in univariable but not multivariable analyses, which may be related to the potentially confounding association between obstructive sleep apnea and hypertension ^56^. We also did not find an association between diabetes mellitus and CVR, which may be a consequence of small sample sizes, nor did we show an association between lipid biomarkers such as LDL-cholesterol and CVR. Indeed, the associations between diabetes mellitus and hyperlipidemia have been less consistently demonstrated in previous studies ^9,17–19^ in accordance with our findings.

The majority of previous studies that have evaluated CVR using fMRI have used 3T MRI and have most commonly acquired CVR data using BOLD labelling ^6^. Indeed CVR values measured using fMRI correlate well with those obtained using other modalities such as positron emission tomography and single-photon emission computed tomography, which require ionizing radiation, and transcranial Doppler, which only evaluates large conductance blood vessels as opposed to the whole-brain vasculature ^6^ like fMRI. When inducing vasodilation, breathing modulation techniques such as breath-holding are commonly used but less often than fixed inspired gas administration or computer-controlled devices ^6^. These latter techniques offer the potential for greater control over the vasodilating stimulus, but may be more cumbersome, may be more difficult for patients to tolerate, and require additional resources. Further, reports have indicated that CVR measures obtained using breath-holding yield results with similar reliability ^6^.

### Limitations

The current study has some limitations. First, our sample is made up of patients who were referred to a tertiary referral center and so comprise a select population. In a similar vein, study participants were referred for a fMRI for clinical reasons related to a diagnosis of epilepsy, and although they did not have baseline CVD, they were not entirely healthy and free of disease. However, few studies linked epilepsy to endothelial dysfunction, suggesting that it did not impose a major interference with our results ^57,58^. Second, we are limited by a cross-sectional design and a relatively small sample size. Thus, we cannot infer causality and larger prospective studies will be required to confirm our findings and to disentangle the direction of association amongst the variables studied. Third, we did not evaluate CVR across separate regions of the brain, which may have been of interest to implicate specific regions in cardiovascular risk prediction. Fourth, typically for cerebrovascular disease absolute quantification of CVR is used but this requires controlled exogenous delivery of CO2 or at least measurement of end tidal pCO2, since units that are typically used are %BOLD signal change/mm Hg (pCO2) rather than T values that merely reflect changes in BOLD signal in the hypercapnic state relative to a normocapnic state. However, as we are considering global average values, it may not make much of a difference since patients with severe carotid steno-occlusive disease, large prior infarcts or cerebral mass lesions were not included and so we can make the assumption that the pCO2 changes across subjects are equivalent. Last, we did not follow the patients prospectively for incident CVD or events which would be an essential next step when determining the clinical utility of measuring CVR when estimating risk in apparently low-risk populations.

## CONCLUSION

Traditional cardiovascular risk factors are significantly associated with abnormal CVR in young adults without established CVD or cerebrovascular disease undergoing fMRI for reasons related to a diagnosis of epilepsy. CVR using fMRI could provide an integrated index of the collective burden of cardiovascular risk factors on the cerebrovascular bed that could be followed across time and could form a therapeutic target to prevent CVD and cerebrovascular events.

## ABBREVIATIONS

BMI: Body Mass Index
BOLD: Blood Oxygen Level Dependent
CAD: Coronary Artery Disease
CBF: Cerebral Blood Flow
CVD: Cardiovascular Disease
CVR: Cerebrovascular Reactivity
fMRI: Functional Magnetic Resonace Imaging
HDL: High Density Lipoprotein
LDL: Low Density Lipoprotein
MI: Myocardial Infarction
TSH: Thyroid Stimulating Hormone

## Data Availability

All data is available for review upon request

